# A Mathematical Model Simulating the Adaptive Immune Response in Various Vaccines and Vaccination Strategies

**DOI:** 10.1101/2023.10.05.23296578

**Authors:** Zhaobin Xu, Jian Song, Hongmei Zhang, Zhenlin Wei, Dongqing Wei, Jacques Demongeot

**Author notes:** Correspondence (Z.X.); (J.D.).

## Abstract

Vaccination is the most effective measure for preventing infectious diseases. Developing an appropriate mathematical model facilitates quantitative research into the activation of adaptive immune responses in the human body by vaccines, thereby providing better guidance for vaccine development. In this study, we have constructed a novel mathematical model to simulate the dynamics of antibody levels following vaccination. Based on principles from immunology, our model provides a concise and accurate representation of the kinetics of antibody response. We have compared the antibody dynamics within the body after administering several common vaccines, including traditional inactivated vaccines, mRNA vaccines, and future attenuated vaccines based on defective interfering viral particles (DVG). Our model explains the crucial role of booster shots in enhancing IgG antibody levels and provides a detailed discussion on the advantages and disadvantages of different vaccine types. From a mathematical standpoint, our model systematically proposes four essential approaches to guide vaccine design: enhancing antigenic T-cell immunogenicity, directing the production of high-affinity antibodies, reducing the rate of IgG decay, and lowering the peak level of vaccine antigen-antibody complexes. Our model contributes to the understanding of vaccine design and its application by explaining various phenomena and providing positive guidance in comprehending the interactions between antibodies and antigenic substances during the immune process.

## 1. Introduction

Vaccination plays a vital role in maintaining human health. The growth of population and increased life expectancy are closely associated with vaccination [1-3]. Many historically significant infectious diseases such as smallpox, plague, and cholera have been completely eradicated or effectively controlled through vaccination [4-6]. However, the emergence of new infectious diseases has challenged the traditional understanding of vaccines. For instance, the appearance of HIV/AIDS made the scientific community realize that not all viral infections can be swiftly addressed with effective vaccines [7-8]. The emergence of the COVID-19 virus has also led people to gradually understand that vaccination does not guarantee lifelong protection, thus challenging the classical theory of herd immunity [9-11].

Modern immunological research methods have made remarkable advancements, with experimental immunology continuously revealing new immune regulatory genes and signaling pathways at the cellular and molecular levels [12-13]. However, these new developments and discoveries often remain fragmented and fail to provide a systematic explanation for the macroscopic manifestations observed in human immune processes, such as differences in the duration of vaccine protection. Mathematical models serve as a valuable tool to address this limitation by enabling systematic and quantitative studies of host-pathogen interactions, thereby more effectively elucidating the mechanisms and progression of infectious diseases. In recent years, mathematical modeling of host-pathogen interactions has provided strong theoretical guidance for the prevention and treatment of infectious diseases [14-18]. This has further led to the development of mathematical modeling in the field of vaccines. For example, Rajat Desikan et al. established vaccine models to predict guidelines for updating vaccines against evolving pathogens like SARS-CoV-2 and influenza in the context of pre-existing immunity [19]. Cristina Leon et al. successfully simulated the innate and adaptive immune responses of hosts to COVID-19 infection or vaccination using a mathematical model [20]. Indrajit Ghosh utilized a mathematical model to study the efficacy of antiviral drugs and vaccination on the dynamics of SARS-CoV-2 infection [21].

Based on the comprehensive review of the aforementioned research, we have advanced the existing antibody kinetics model [22] by incorporating a novel component—vaccination. Our focus lies in investigating the specific activation mechanisms of vaccines on host adaptive immune responses. This paper presents a meticulous examination of our research findings, starting with a systematic overview of our model and its distinguishing features compared to other models. Within this context, we establish a clear delineation of various parameters within the model, attributing them to factors such as viral pathogenicity, clinical symptom severity, and antigen-specific T-cell immunogenicity.

Subsequently, employing our refined model, we extensively evaluate the performance of distinct vaccines and varied administration strategies. Moreover, utilizing the developed model as a foundation, we propose four fundamental strategies to guide vaccine design: augmentation of antigen-specific T-cell immunogenicity, targeted elicitation of high-affinity antibodies, attenuation of IgG decay rate, and reduction of peak levels in vaccine antigen-antibody complexes. Our model provides a comprehensive and quantitative elucidation of the inducible effects of diverse antigenic substances on adaptive humoral immunity. Consequently, it offers valuable theoretical insights for future endeavors in mathematical modeling and experimental investigations in the field.

## 2. Materials and Methods

### 2.1. An overview of the immnodynamic model

Before delving into the specific mathematical equations, we initially present a macroscopic overview of our model to enhance the reader’s comprehension of this mathematical framework. Our model can be succinctly represented by the aforementioned flowchart, which encompasses five core components and thirteen significant reactions. Specifically, Reaction 1 denotes the binding of B cells producing IgM with antigenic substances, resulting in the generation of antigen-antibody complexes. Simultaneously, these antigen-antibody complexes interact with Th cells, eliciting immunological effects on Th cells. The antigenic substances implicated in this reaction may encompass protein constituents found in inactivated vaccines, live viruses, or those translated from mRNA vaccines. Reaction 2 signifies the recognition and swift elimination of IgM-antigen complexes by the immune system, potentially involving various immune cells, including NK cells.

Reaction 3 embodies the proliferative influence of Th cells on adjacent B cells, a process that is frequently overlooked in prevailing mathematical models. We explicitly incorporate this positive feedback effect into our model through Reaction 3. When B cells expressing specific antibodies bind to antigens, the antigens are recognized, engulfed, and lysed by B cells, leading to the production of corresponding peptide fragments. These peptide fragments possess distinct Th cell immune stimulatory properties. The binding between B cells and Th cells is facilitated via the interaction between B cell antibodies and Th cell binding sites, which may encompass intermediate molecules such as CD8. The lysed peptide fragments are presented to Th cells, triggering alterations in their signal transduction pathways and subsequent secretion of diverse cytokines. These cytokines facilitate the proliferation of themselves and neighboring cells. Consequently, B cells recognizing the relevant antigen and their corresponding Th cells can experience rapid and substantial proliferation within a short timeframe [26-27]. Thus, we posit that the regeneration of B cells, or more precisely, the regeneration of antibodies, emanates from antigen-antibody complexes. Hence, the rate of regeneration is directly proportional to the concentration of antigen-antibody complexes.

Reaction 4 embodies the process by which IgM transforms into IgG. There are two sources of IgG-producing B cells: one stems from the transformation of IgM of the same isotype, while the other emerges through the proliferation of B cells themselves, a process denoted as Process 10. This transformation is often neglected in prevailing models, which fail to consider the relationship between IgM and IgG. This omission detrimentally impacts the accuracy and reliability of the models. For instance, dengue virus infection models invariably necessitate dynamic simulations involving IgM. However, models that account for IgM often isolate it from IgG for separate analyses, which evidently lacks veracity. We shall demonstrate the profound implications of the IgM-IgG relationship in the subsequent results, wherein the selection of vaccination strategies is concerned. Notably, many vaccines employ booster doses precisely due to this relationship. IgM belongs to the initial class of antibodies within the human body, boasting an antibody repertoire significantly larger than that of IgG. For viruses to which we have never been exposed, our IgM antibody repertoire comprises antibodies with robust binding affinity, while the IgG antibody repertoire may lack antibodies of comparable strength. Accordingly, when faced with such infections or receiving the initial dose of such vaccines, neutralizing antibodies derive from IgM, whereas IgG production occurs subsequently, initially stemming from the transformation of IgM.

Reaction 5 denotes the binding of self-antigenic substances to IgM. Our previous antibody kinetics theory has substantiated the role of environmental or self-antigens in preserving antibodies. This holds true for both IgM and IgG. Reaction 6 represents the clearance of IgM-self-antigen complexes, whereas Reaction 7 illustrates the stimulating effect of IgM-self-antigen complexes on IgM regeneration. Considering the role of self-antigens in antibody preservation constitutes another pivotal characteristic of our model. The neglect of self-antigenic substances would result in degradation, and subsequently to the loss of the initial antigenic stimulus, all antibodies would steadily decline. This would inevitably lead to the eradication of antibodies. Consequently, numerous models fail to explicate the phenomenon of long-term antibody persistence and the provision of sustained protection. Hence, we introduce self-antigenic substances. In fact, self-antigens play an extremely crucial role in the maturation, differentiation, and clonal selection of immune cells. We have conducted more comprehensive research on this subject matter in a previous article [28]. The same principle applies to the maintenance of IgG by self-antigens, represented by Reactions 11, 12, and 13. Concerning IgG, the interaction between antigens and IgG is the same type as that between IgM and antigens, and it is delineated by Reactions 8, 9, and 10.

### 2.2. A mathematical modeling of adaptive immune response in different vaccine types

According to Figure 1, we have listed the reactions involved and presented them in Table 1. Table 2 and Table 3 represent the selection of parameters and the values of initial variables, respectively. The antigen replication reaction is represented as reaction 14. All degradation processes are represented as reaction 15-19.

**Table 1:** Reaction index and the name of each reaction in our mathematical model.

| Reaction index | Reaction |
| --- | --- |
| 1 | $\text{IgM} + \text{Antigen} \xrightleftharpoons[k_{-1}]{k_1} \text{IgM-Antigen complex}$ |
| 2 | $\text{IgM-Antigen complex} \xrightarrow{k_2} \text{Removed by immune system}$ |
| 3 | $\text{IgM-Antigen complex} \xrightarrow{k_3} \text{IgM}$ |
| 4 | $\text{IgM} \xrightarrow{k_4} \text{IgG}$ |
| 5 | $\text{IgM} + \text{Self-Antigen} \xrightleftharpoons[k_{-5}]{k_5} \text{IgM-Self-Antigen complex}$ |
| 6 | $\text{IgM-Self-Antigen complex} \xrightarrow{k_6} \text{Removed by immune system}$ |
| 7 | $\text{IgM-Self-Antigen complex} \xrightarrow{k_7} \text{IgM}$ |
| 8 | $\text{IgG} + \text{Antigen} \xrightleftharpoons[k_{-8}]{k_8} \text{IgG-Antigen complex}$ |
| 9 | IgG-Antigen complex $\xrightarrow{\quad\quad\quad}$ Removed by immune system |
| 10 | IgG-Antigen complex $\xrightarrow{k_{10}}$ IgG |
| 11 | IgG + Self-Antigen $\xrightleftharpoons[k_{-11}]{k_{11}}$ IgG-Self-Antigen complex |
| 12 | IgG-Self-Antigen complex $\xrightarrow{k_{12}}$ Removed by immune system |
| 13 | IgG-Self-Antigen complex $\xrightarrow{k_{13}}$ IgG |
| 14 | Antigen $\xrightarrow{k_{14}}$ Antigen |
| 15 | IgM $\xrightarrow{k_{15}}$ Degradation |
| 16 | IgG $\xrightarrow{k_{16}}$ Degradation |
| 17 | Antigen $\xrightarrow{k_{17}}$ Degradation |
| 18 | mRNA $\xrightarrow{k_{18}}$ Antigen |
| 19 | mRNA $\xrightarrow{k_{19}}$ Degradation |

**Table 2:**
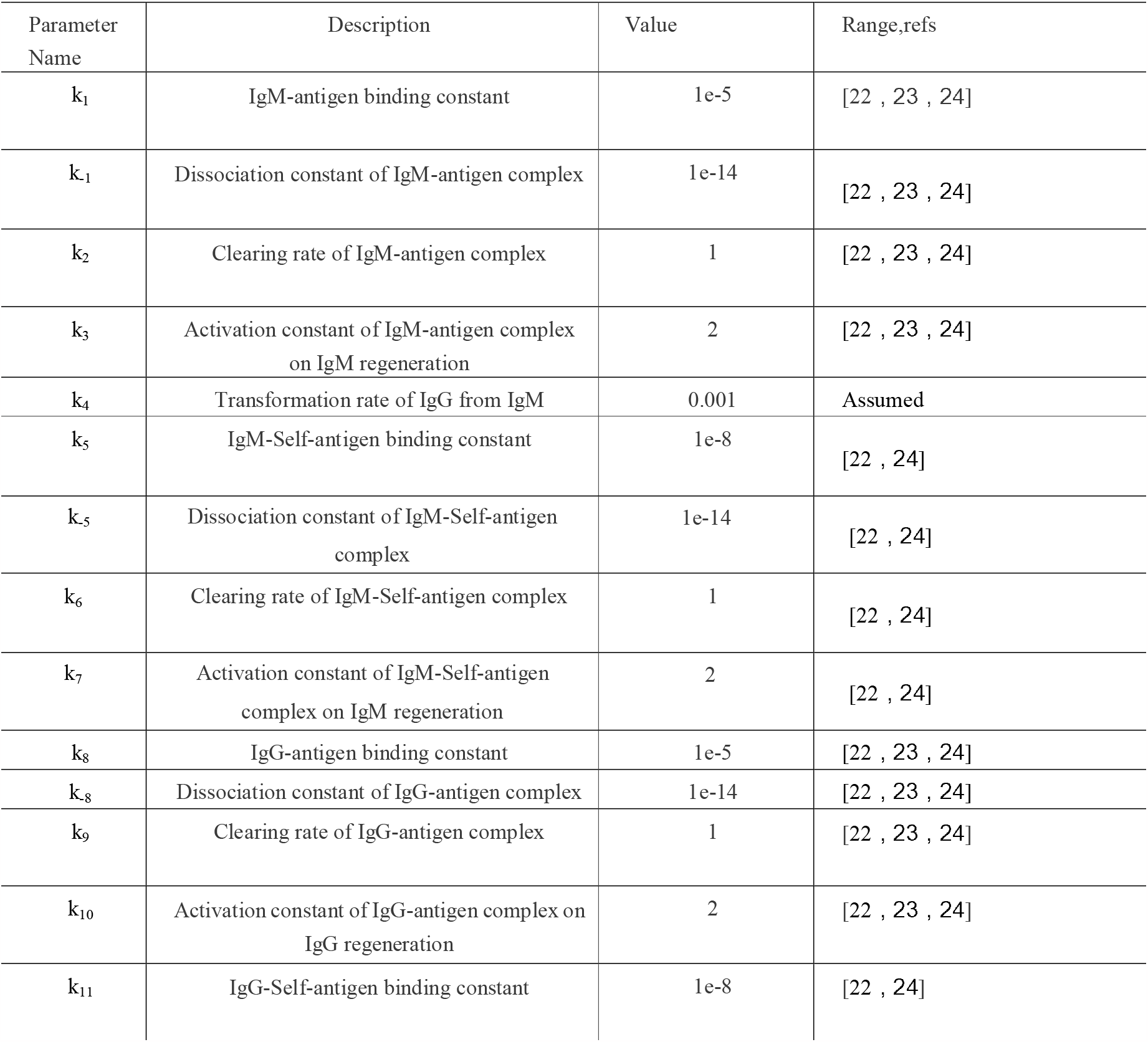
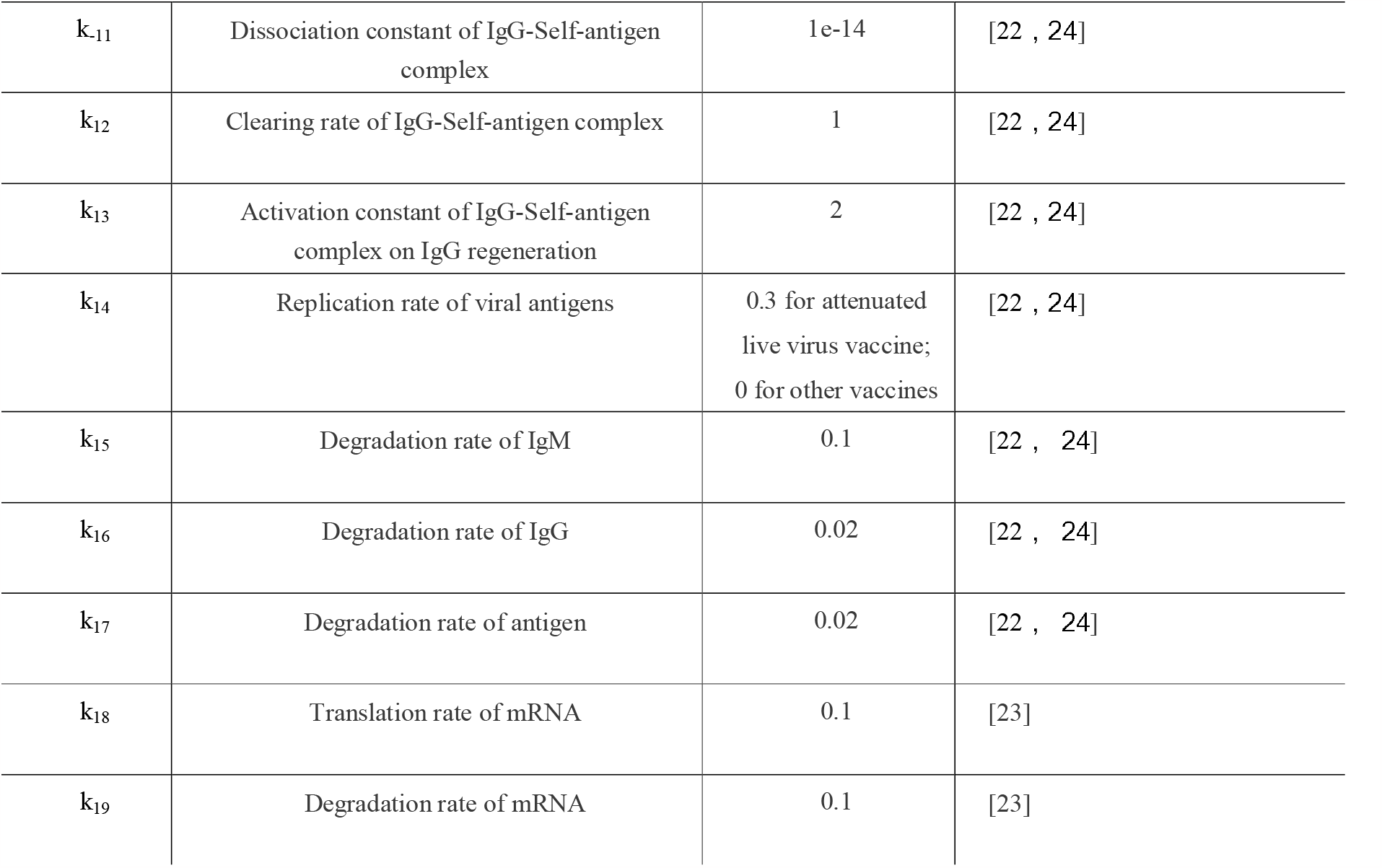
Estimates of the calibrated model parameters.

**Table 3:**
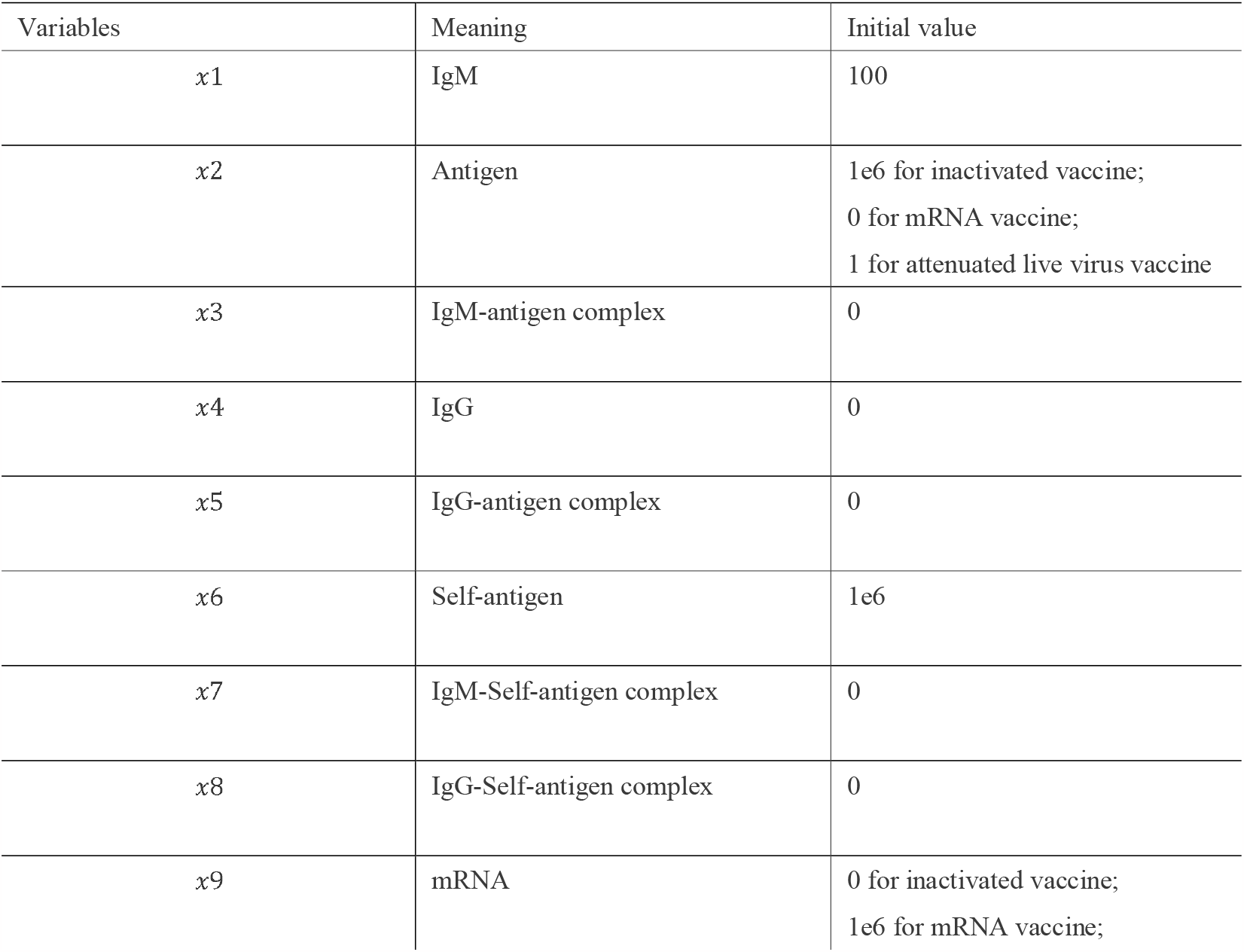
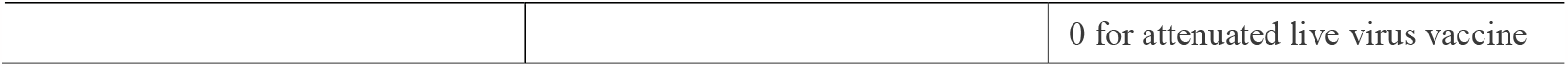
Time-dependent variables of the mathematical model characterizing the antibody-antigen interactions.

| Variables | Meaning | Initial value |
| --- | --- | --- |
| $x_1$ | IgM | 100 |
| $x_2$ | Antigen | 1e6 for inactivated vaccine;<br>0 for mRNA vaccine;<br>1 for attenuated live virus vaccine |
| $x_3$ | IgM-antigen complex | 0 |
| $x_4$ | IgG | 0 |
| $x_5$ | IgG-antigen complex | 0 |
| $x_6$ | Self-antigen | 1e6 |
| $x_7$ | IgM-Self-antigen complex | 0 |
| $x_8$ | IgG-Self-antigen complex | 0 |
| $x_9$ | mRNA | 0 for inactivated vaccine;<br>1e6 for mRNA vaccine; |
|  |  | 0 for attenuated live virus vaccine |

**Figure 1:**
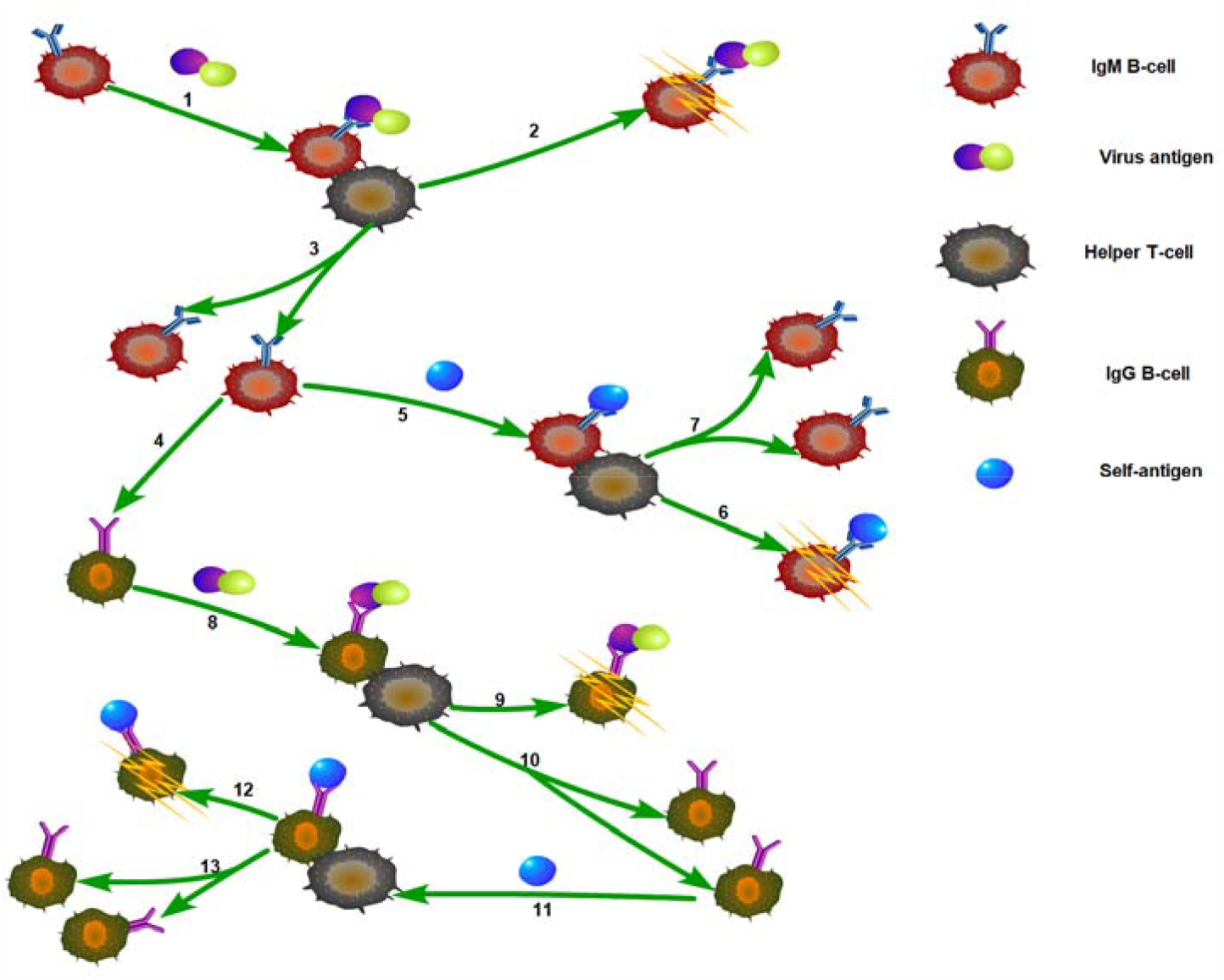
Model schematic. 13 core reactions and 5 components are illustrated in the model. Those 5 components include virus antigen, self-antigen, IgM-producing B-cell, IgG-producing B-cell, and helper T-cell. Each reaction is represented in green line with arrow indicating the reaction direction.

Corresponding ordinary differential equations (ODEs) can be derived as follows:

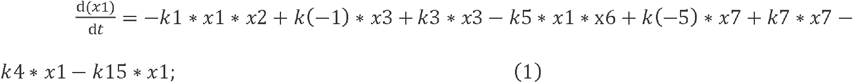

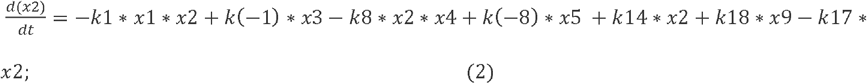

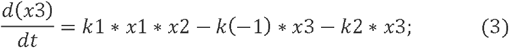

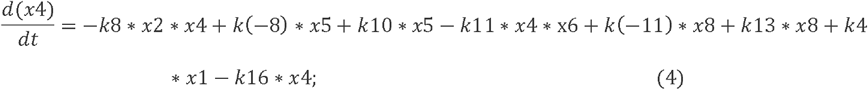

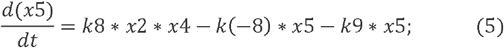

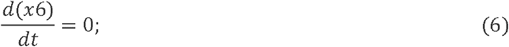

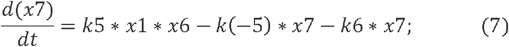

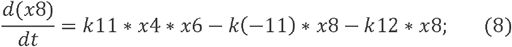

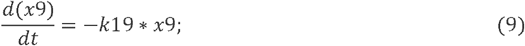

The above equations were solved numerically using the ode15s function in MATLAB [25]. It is important to note that the concentration of self-antigen-like substances is considered constant, as they can be rapidly replenished from the environment. Therefore, the sixth term in the equation is set to 0.

For inactivated vaccines, such as inactivated primary vaccines or inactivated recombinant vaccines like adenovirus vector vaccines, the viruses are unable to replicate. Consequently, the value of k14 is set to 0. Similarly, for mRNA vaccines, the expressed protein antigens cannot undergo self-amplification, resulting in k14 also being equal to 0. However, in the case of attenuated vaccines, this value is non-zero. Unlike regular viruses, attenuated vaccines, such as those based on defective viruses, exhibit significantly reduced replication activity. Thus, the value of k14 is significantly smaller compared to regular viral infections.

For mRNA vaccines, mRNA molecules can be transcribed into antigen-like substances. Therefore, the initial concentration of antigen-like substances is set to 0, while the initial mRNA concentration is assigned a higher value. Conversely, for inactivated and attenuated vaccines, the initial mRNA concentration is assumed to be 0. Although attenuated viruses still rely on mRNA for replication, we have omitted the consideration of mRNA in the model. The ultimate outcome of virus replication is the production of replicated antigens.

Another notable distinction between inactivated vaccines and mildly pathogenic virus infections lies in the difference in initial antigen concentration. Inactivated vaccines, due to the inability of antigens to self-replicate, require a higher injection concentration to achieve a desirable immune response. Conversely, there is no dosage threshold for vaccines against mildly pathogenic virus infections. As a result, the initial invading viral antigen concentration can be relatively low, yet still elicit an effective immune response.

## 3. Results

### 3.1. Kinetic Modeling of Antibody Response Following Inactivated Vaccine Administration

Inactivated vaccines are a conventional method of vaccine production. The term “inactivated vaccines” includes the traditional approach of rendering the original pathogens non-infectious. However, it also encompasses any method of antigen delivery that utilizes non-replicating forms of antigens. This definition extends to include genetically modified inactivated vaccines, including those created through gene recombinant techniques. Nevertheless, conventional inactivated vaccines face significant challenges when it comes to preparing RNA virus vaccines. A major obstacle is the instability of RNA viruses during replication cycles, leading to an increasing proportion of defective viral genomes (DVGs) with each passage. Beyond a particular generation, this results in the total loss of viral replication activity [29]. However, genetically engineered vaccines have been industrially developed for preventing RNA virus infections, such as the AstraZeneca vaccine for COVID-19 [30]. Booster doses are often required to achieve optimal efficacy for most inactivated vaccines. Our simulation elucidates the underlying mechanisms involved.

As illustrated in Figure 2, solid lines represent concentration changes after the initial dose, while dashed lines represent concentration changes of various substances after the second dose. Maintaining elevated levels of IgG within the body is crucial for preventing reinfection due to its significantly slower decay rate compared to IgM. From Figure 2, it can be observed that the initial increase in IgG levels following the first dose is limited (represented by the green solid line), while there is a substantial increase in IgM levels (represented by the red solid line). This is primarily attributed to the initial scarcity of IgG within the body at the time of vaccination. Consequently, a small amount of IgG is derived from the conversion of IgM, accompanied by self-amplification of IgG through antigen binding. Simultaneously, IgM undergoes rapid proliferation, followed by swift decay once antigen-like substances are depleted. IgM ceases proliferation when it is no longer stimulated by antigens, resulting in rapid decay. A fraction of IgM continues transforming into IgG during this process.

**Figure 2:**
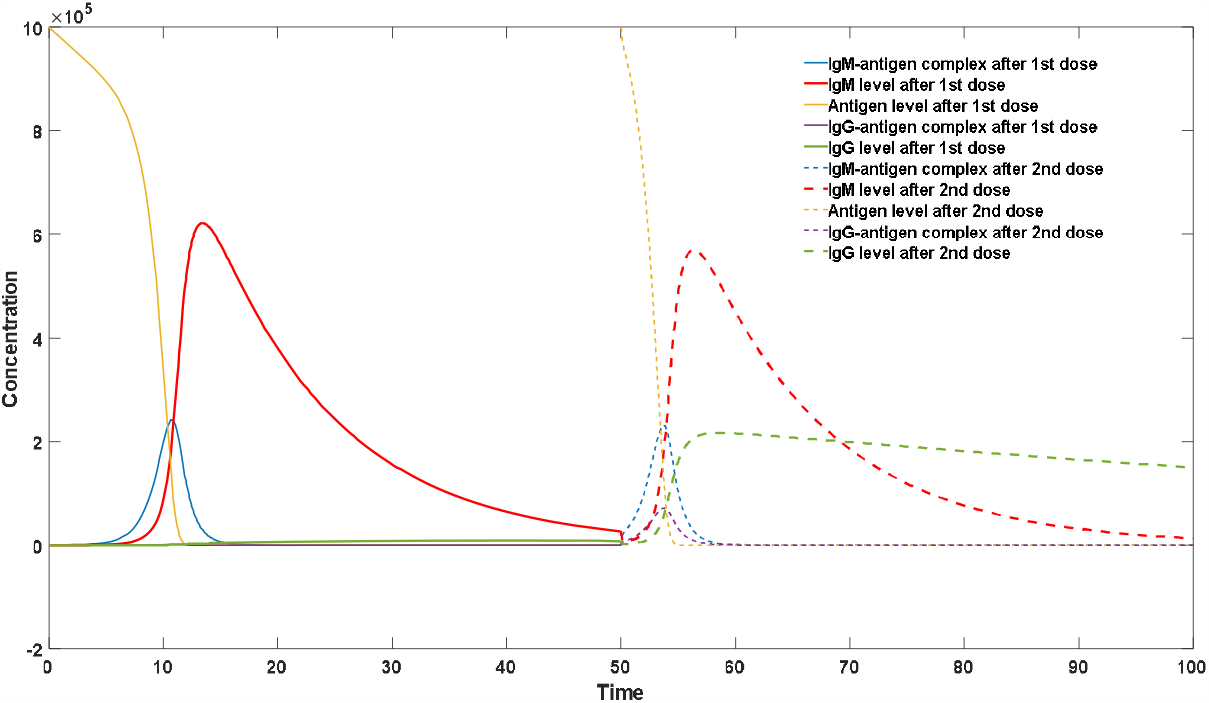
Antibody dynamics after 2 doses of inactivated vaccines. First dose is injected at the initial time unit, second dose is injected at 50^th^ time unit. Both injection dosages of antigen substances are 10^6^.

At 50^th^ time units, the second dose is administered, resulting in a significantly different pattern. IgG experiences rapid growth, with its concentration quickly rising to a higher level (>2e5). Correspondingly, IgM levels also increase (represented by the red dashed line). This phenomenon stems from the presence of a certain level of IgG attained after the initial vaccination. The substantial increase in IgG content during the second dose primarily arises from the proliferation process of IgG itself, stimulated by antigens, as described in the relevant reaction (Reaction 10) outlined in the methods section. Administering booster doses allows IgG levels to reach a considerable threshold, thereby extending the protective effect and duration beyond that achieved by a single dose. This is why multiple-dose administration strategies are commonly employed for vaccines [31]. Notably, in Figure 2, we not only describe the kinetic changes of antibodies but also specifically depict the dynamics of IgG-antigen and IgM-antigen complexes. These antigen-antibody complexes play a critical role in the immune response. They not only participate directly in antibody regeneration through feedback regulation but also serve as a direct indicator of the intensity of patient symptoms. Symptoms resulting from viral infections or vaccine administration, such as fever, are positively correlated with the concentration of antigen-antibody complexes rather than the concentrations of viruses or antibodies alone. Therefore, when evaluating vaccine side effects, it is necessary to consider changes in the concentration of antigen-antibody complexes induced by the vaccine. Figure 2 shows that the blue curve represents the concentration changes of IgM-antigen complexes, with their peaks exceeding 2e5 in both doses, while the concentration of IgG-antigen complexes also exhibits a noticeable increase after the second dose (represented by the purple dashed line). Additionally, inactivated vaccines may have a potential drawback when antigen structures may undergo changes after treatment with high temperatures or chemical reagents. Such alterations can result in modifications to antigenic determinants, similar to the effect of antigenic drift, potentially leading to a significant decline in vaccine efficacy [32-33].

### 3.2. Kinetic Modeling of Antibody Response Following mRNA Vaccine Administration

With increasing understanding of immunological mechanisms and advancements in mRNA preparation and packaging technologies, mRNA vaccines have made significant developments in recent years. Pfizer’s BNT162b2 and Moderna’s mRNA-1273 are a few successful examples that have shown better protective efficacy compared to traditional vaccines in clinical trials [34-35]. The advantages of mRNA vaccines lie in the preservation of the original antigen structure due to the lack of inactivation processes, which reduces the likelihood of antigenic drift [36]. Adverse reactions are often lower for mRNA vaccines due to the gradual process of antibody synthesis upon mRNA entry into the body, resulting in lower concentrations of antigen-antibody complexes observed in the model. Specifically, the blue curve in Figure 3 represents the concentration changes of IgM-antigen complexes. It can be seen from Figure 3 that the peak concentrations of IgM-antigen complexes induced by the two doses are significantly smaller compared to those of traditional inactivated vaccines. Similarly to inactivated vaccines, the IgG levels after two doses of mRNA vaccines are significantly higher than those after a single dose, indicating that multiple doses remain the optimal vaccination strategy for mRNA vaccines. It is worth noting that both the final IgG concentration of inactivated vaccines and mRNA vaccines are closely related to the vaccination dose. Excessive vaccination doses can lead to the formation of an excessive amount of antigen-antibody complexes, resulting in severe vaccine side effects. Insufficient vaccination doses are not enough to induce IgG to reach a sufficient concentration level, leading to decreased protection efficiency and shortened protection duration. The optimal vaccination dose can be determined more scientifically through mathematical models. However, it should be noted that both inactivated vaccines and mRNA vaccines require a certain vaccination dose, which poses short-term production obstacles and constraints on their promotion. For highly contagious respiratory system viruses, surpassing the rate of natural infection with vaccine usage is crucial for disease prevention, which constrains the future development of vaccines [37-38]. Due to mRNA’s rapid degradation rate compared to proteins, mRNA vaccines also have the disadvantage of being difficult to store, which brings many inconveniences during usage [39]. Another fatal flaw is the potential for myocarditis. mRNA must be encapsulated in carriers to enter cells for the translation of corresponding antigenic substances. These carriers, different from the spike protein of the original virus, can indiscriminately infect all cells, including non-epithelial tissues and cells with extremely low ACE2 receptor expression. This can lead to potential infection of cardiomyocytes, and during the process of clearing infected cells after antibody production, it can cause damage to cardiomyocytes [40].

**Figure 3:**
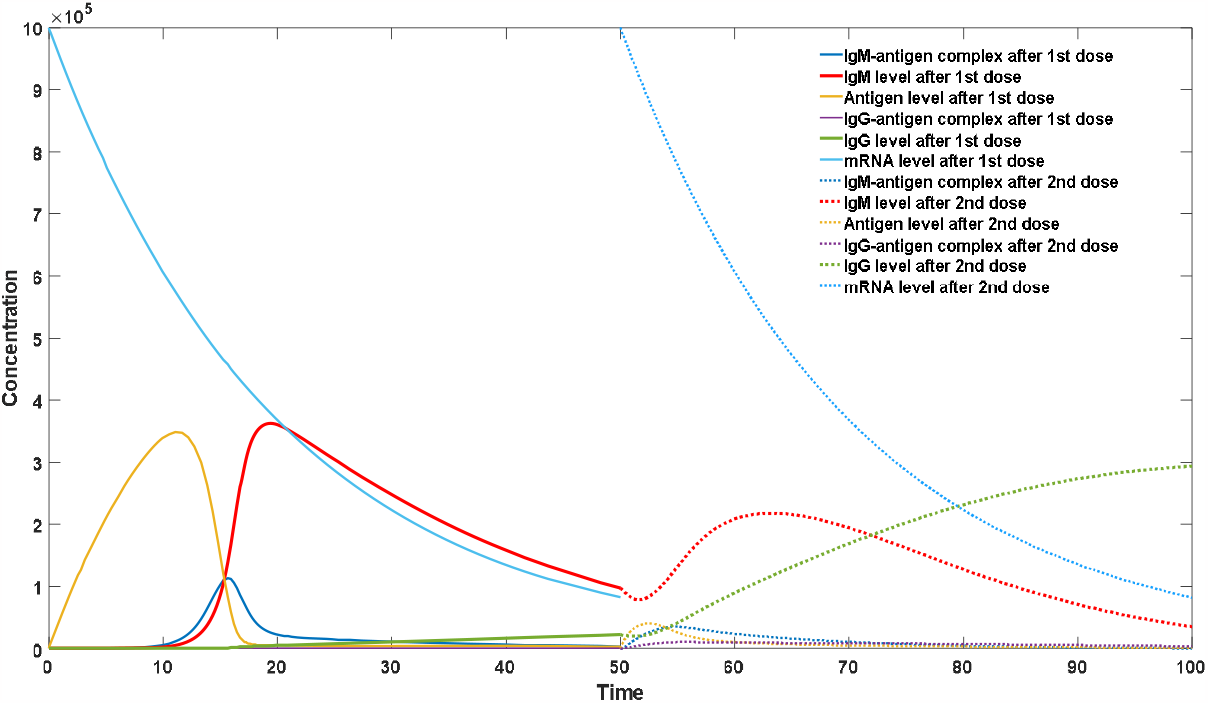
Antibody dynamics after 2 doses of mRNA vaccines. First dose is injected at the initial time unit, second dose is injected at 50^th^ time unit. Both injection dosages of mRNA are 10^6^.

### 3.3. Kinetic Modeling of Antibody Response Following Attentuated Vaccine Administration

The concept of using attenuated vaccines has recently been proposed. Attenuated vaccines refer to the immunization method in which hosts are infected with viruses that have weakened replicability, thereby stimulating their antibody-mediated immunity. As our understanding of viruses deepens, particularly regarding defective viruses [41-43], it is increasingly recognized that a single viral strain may possess various defective genotypes, leading to a decrease in replication activity and milder symptoms. In the case of COVID-19 infection, many asymptomatic cases result from infections with defective viruses. Therefore, some scholars have proposed and put into practice the use of attenuated viruses as natural vaccines [44]. There are various means to reduce viral activity, such as using rare codons in the host to reduce translation efficiency [45-46] or utilizing defective viruses [44,47]. The characteristics of antibody response induced by attenuated vaccines are illustrated in Figure 4. From Figure 4, it can be observed that the IgG levels after attenuated vaccine administration exhibit a significant increase (indicated by the solid green line), surpassing the effects of mRNA and inactivated vaccines after two doses. Meanwhile, it is noteworthy that the immune response triggered by attenuated vaccines is not overly intense, as reflected by relatively low levels of antigen-antibody complexes. The peak concentration of IgG-antigen complexes (indicated by the dashed purple line) is around 2.*5e5, while the peak concentration of IgM-antigen complexes (indicated by the dashed blue line) is around* 1.5e5. The advantages of attenuated vaccines can be summarized as follows: firstly, a high level of IgG can be achieved without the need for multiple doses. Unlike traditional vaccine administration, the antibody response induced by attenuated vaccines exhibits kinetics similar to those resulting from actual viral infections. This is due to the low viral inoculum, which allows sufficient time for viral replication, enabling the conversion from IgM to IgG. Hence, a significant increase in IgG levels can often be achieved with a single low-dose infection caused by attenuated viruses. Secondly, attenuated vaccines require a minimal amount of vaccine dose. Due to the inherent replicability of the virus, only a small amount of attenuated virus is required to stimulate an adequate level of antibodies, and the final antibody levels are not significantly correlated with the vaccine dose. This greatly reduces production costs and usage cycles. Thirdly, attenuated vaccines exhibit good transmissibility. As attenuated vaccines are essentially live viruses, they possess similar infectivity to the original virus. Consequently, unvaccinated individuals can acquire the virus from vaccinated individuals, further increasing the rate of achieving herd immunity. Lastly, attenuated vaccines closely resemble the original virus, possessing antigenic epitopes that are more representative of the original virus and a diverse range of antigenic epitopes. This results in a superior immune response. Although mRNA vaccines have demonstrated good antigenic determinants, their expression primarily focuses on specific antigens rather than the entire virus. Specific antigens often exist in a free state, exposing fewer antigenic epitopes that are absent in their natural conformation. Antibodies induced by such epitopes are evidently unable to provide protection against real infections. However, it should be noted that the development of attenuated vaccines is still in its early stages. These vaccines may pose significant risks to individuals with compromised immune function. The design of attenuated vaccines requires strict control over viral replicability, as excessive replication activity can lead to severe side effects, while insufficient replication activity may fail to stimulate an adequate concentration of protective antibodies.

**Figure 4:**
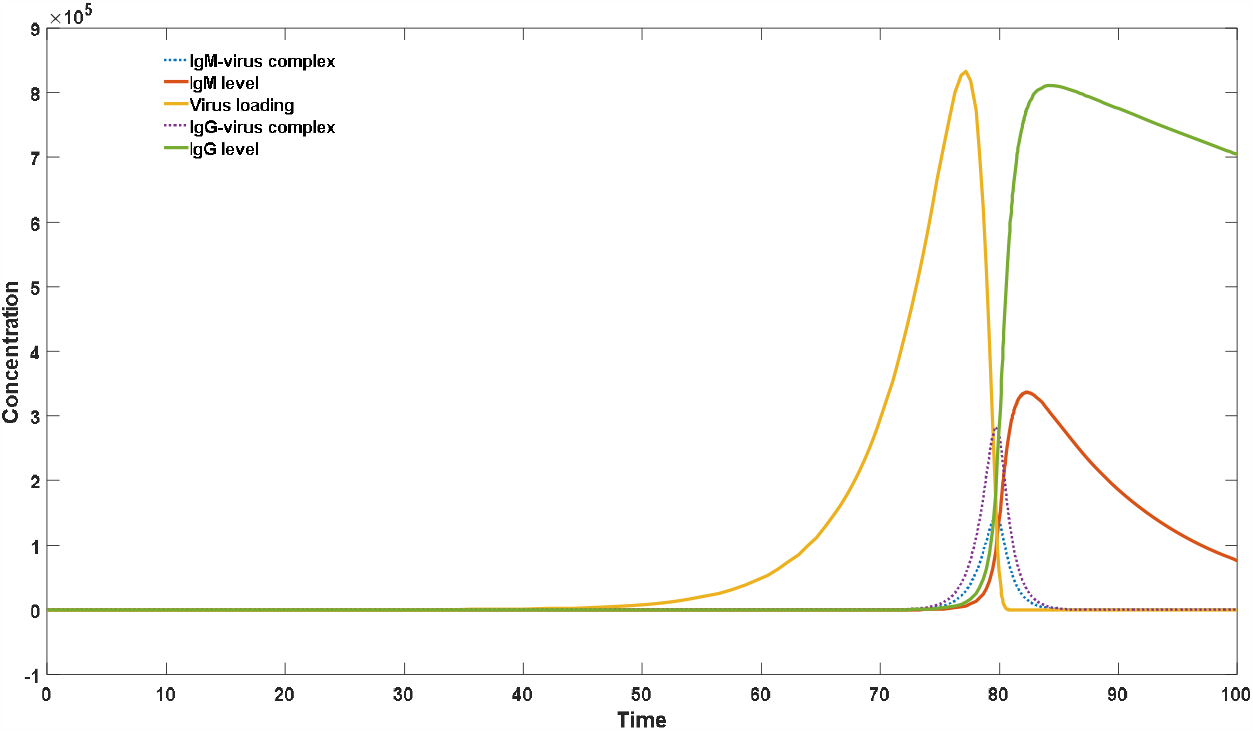
Antibody dynamics after attenuated virus vaccination. Only one injection is implemented at the initial time with an attenuated live virus.

### 3.4. The Impact of Viral Inoculum Dose on the Ratio of IgM to IgG

The proportional relationship between IgM and IgG has been extensively studied in the context of dengue fever virus, where initial attention was drawn to this phenomenon. It has been observed that following primary dengue infection, there is a significant increase in IgM levels, while the rise in IgG levels is not prominent [48-49]. However, during secondary dengue infection, a substantial increase in IgG is observed, akin to the response seen after secondary vaccination. In contrast, for respiratory infectious diseases such as COVID-19, a significant elevation in IgG levels can generally be detected after the initial infection. The discrepancy in these observations can be attributed to differences in viral inoculum dose. A high viral inoculum dose, similar to that of inactivated vaccine administration, rapidly boosts IgM levels but due to the absence of an initial IgG reservoir, the initial IgG levels are negligible. Conversion from IgM to IgG is dependent on the inefficient process of isotype switching, resulting in limited IgG elevation during the initial infection. However, this scenario changes during secondary infections, as explained in detail in section 3.2. For bloodborne infectious diseases like dengue fever, where the viral inoculum dose is higher, the dynamics of IgM and IgG resemble that of vaccine administration. Conversely, for respiratory infectious diseases like COVID-19, where the viral inoculum dose is low, ample time is provided for IgG conversion, extending the incubation period and leading to a higher IgG/IgM ratio post-infection [50]. As shown in Figure 5, an increase in viral inoculum dose from 1 to 100, compared to Figure 4, results in a shorter incubation period and a significant rise in induced IgM levels (solid red line) along with a marked decrease in IgG levels (solid green line), leading to a highly significant reduction in the IgG/IgM ratio.

**Figure 5:**
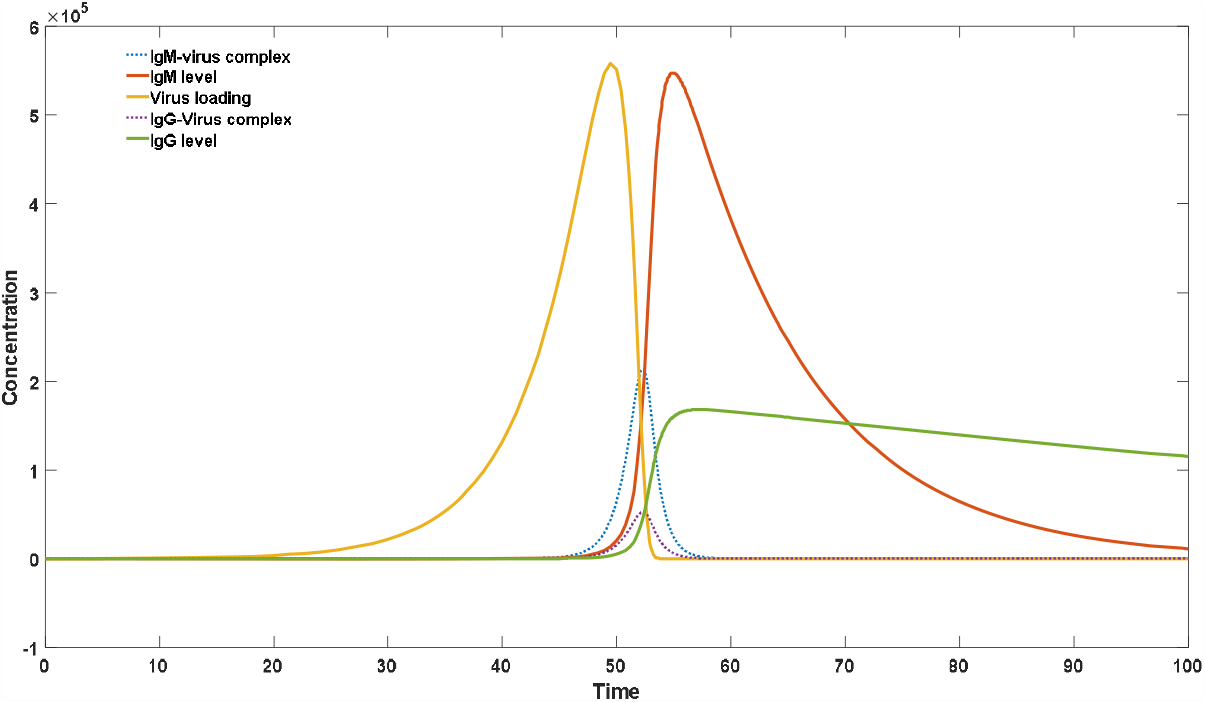
Dosage effect on IgM/IgG ratio. Only one injection is implemented at the initial time with larger number of attenuated live viruses (100 in this case).

### 3.5. Calculation of the Protection Time Brought by Vaccination

The term “herd immunity” was frequently mentioned prior to the large-scale administration of COVID-19 vaccines. When the effective reproduction number (*R*_*0*_) of a virus is known, achieving herd immunity can be accomplished by vaccinating a proportion of the population equal to 1-1/ *R*_*0*_, thereby fundamentally eradicating the infectious disease. For instance, this approach was successfully employed to eliminate diseases with high transmissibility, such as smallpox. However, in the case of combating COVID-19, it has gradually been recognized that in addition to viral mutation effects, antibodies can undergo decay, resulting in time-limited protection. This waning protection implies that for the majority of the population, repeated infections become unavoidable, rendering the theory of herd immunity inapplicable to certain infectious diseases. In our previous work [22], we explained in detail why certain vaccines, such as those for smallpox and mumps, confer lifelong protection, while others, such as the hepatitis B vaccine, provide protection for a period of over a decade, and vaccines like the COVID-19 and influenza vaccines offer shorter-term protection, typically within one year.

Studying the duration of vaccine-induced protection is analogous to studying the duration of naturally acquired immunity, both of which require calculation of the critical threshold levels of IgG. However, it should be noted that the critical thresholds differ for different antibody isotypes, with high-affinity antibodies having much lower thresholds compared to low-affinity antibodies. Once we have determined the host’s antibody kinetic parameters, we can use our model to calculate the duration of vaccine-induced protection. Our calculation methodology is straightforward: we simulate viral invasion at different time points by setting the viral quantity at that specific time to 1. We then observe the dynamic changes in the virus and antibody populations. If significant viral proliferation and peak concentrations of antigen-antibody complexes occur during subsequent time points, it indicates an infection. When the concentration is relatively low, as shown in Figure 6a, the infection may be asymptomatic or mild. When the concentration is higher, as shown in Figure 6b, it corresponds to symptomatic infection. Figures 6a and 6b represent calculated diagrams illustrating the duration of protection for all infections and the duration of protection specifically against symptomatic infections following vaccination, respectively. In Figure 6a, the actual viral invasion occurs at 200 time units, with viral proliferation and antibody elevation observed in the red region on the right after a long incubation period. At this point, the concentration of IgG-antigen complexes increases only slightly, which can be considered a case of asymptomatic infection. The critical time unit, marked as the 200th time unit, is the threshold before which viral infection barely leads to any viral proliferation due to rapid neutralization by high concentrations of IgG. Beyond this critical point, the virus demonstrates varying degrees of proliferation, and the later the invasion occurs, the lower the IgG concentration, resulting in more pronounced viral proliferation. As shown in Figure 6b, when viral invasion occurs at the 400th time unit, significant viral proliferation, antibody elevation, and antigen-antibody complex elevation are observed. The concentration of antigen-antibody complexes exceeds 1e5, which can be considered as the critical concentration for symptomatic infection. Therefore, viral invasions occurring before the 400th time unit do not lead to symptomatic infections, while those occurring thereafter consistently result in symptomatic infections. It can be observed that the duration of protection against symptomatic infections conferred by the vaccine is significantly longer than the duration of protection against all infections. An interesting phenomenon is that after vaccination or natural infection, to prevent the recurrence of severe infections, it is advisable to have moderate exposure to the virus rather than achieving complete self-protection. As shown in Figure 6a, early exposure to the virus without significant symptoms can lead to a re-elevation of IgG antibody levels, thus providing more durable subsequent protection. Complete avoidance of virus exposure may result in more pronounced clinical symptoms upon encountering the virus at a later stage, as demonstrated in Figure 6b. The later the viral invasion occurs, the higher the peak concentration of antigen-antibody complexes, leading to more pronounced symptoms. It is for this reason that we do not recommend long periods of excessive self-protection, such as habitual mask wearing.

**Figure 6a:**
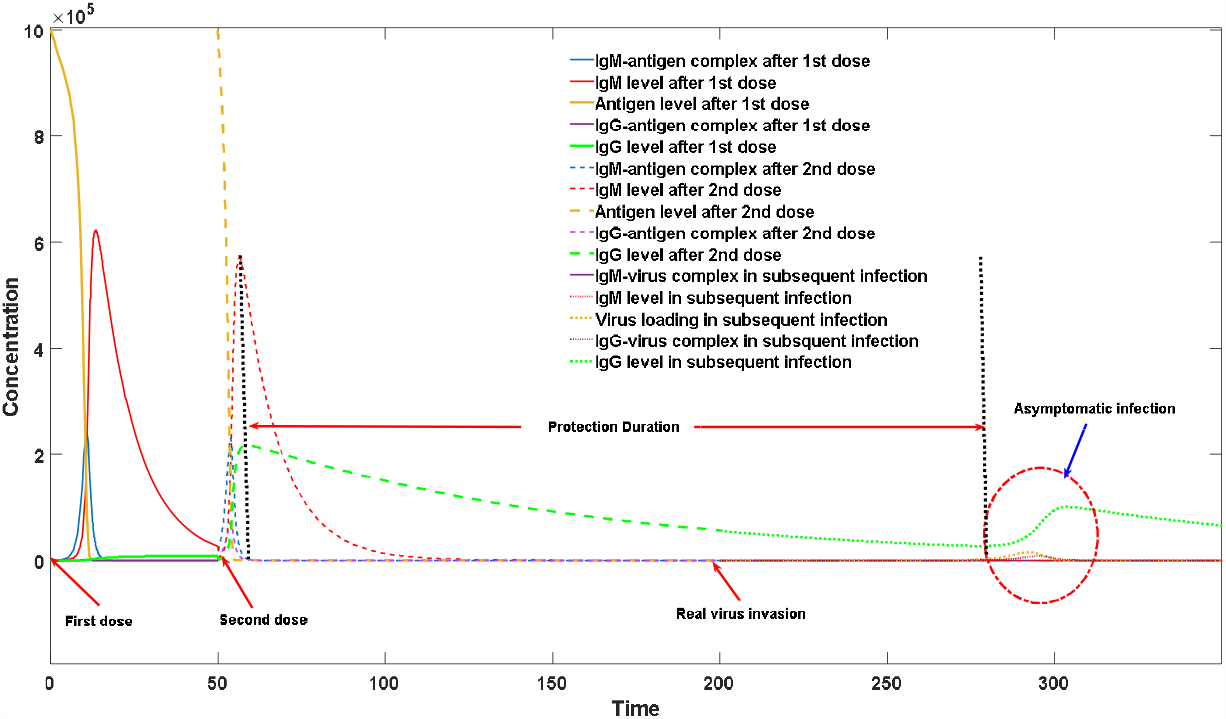
An illustration of protection time calculation toward asymptomatic infection. First dose is injected at the initial time unit, second dose is injected at 50^th^ time unit. Both injection dosages of antigen substances are 10^6^. Single live virus invaded at 200^th^ time unit. The protection duration and subsequent infection curve are both marked in this figure.

**Figure 6b :**
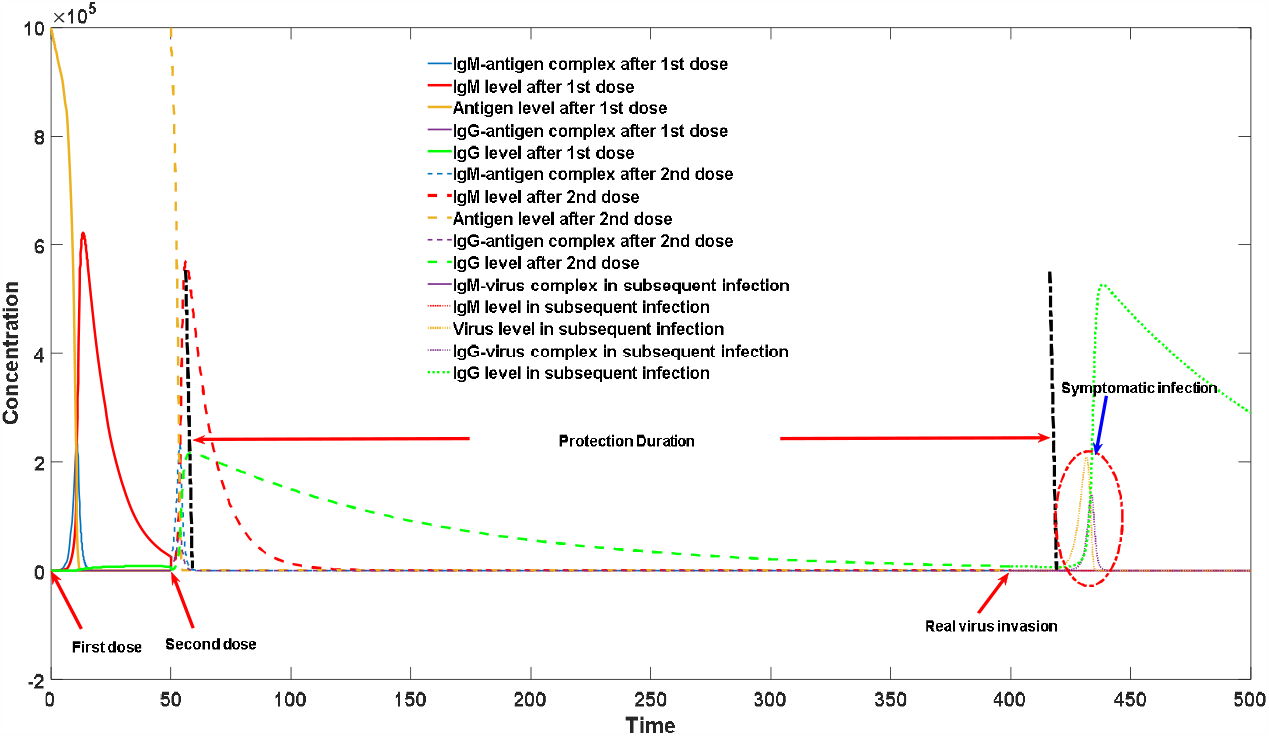
An illustration of protection time calculation toward symptomatic infection. First dose is injected at the initial time unit, second dose is injected at 50^th^ time unit. Both injection dosages of antigen substances are 10^6^. Single live virus invaded at 400^th^ time unit. The protection duration and subsequent infection curve are both marked in this figure.

### 3.6. Four Major Approaches Guiding Vaccine Design

In the preceding sections, we discussed mathematical models for various types of vaccine administration. Based on these models, we now propose four guiding principles for future vaccine design. While lacking systematic overview and theoretical summarization, many of these approaches have been attempted and implemented by scientists.

#### 3.6.1 Enhancing the T-cell Immunogenicity of the Antigen

The T-cell immunogenicity of antigens plays a crucial role in stimulating antibody proliferation, yet it is often overlooked in vaccine design. In fact, the T-cell immunogenicity of antigens forms the basis for host recognition of self and foreign components. When antibodies generated within the body strongly bind to self-antigenic substances due to the low T-cell immunogenicity of self-antigens, they cannot undergo extensive proliferation with the assistance of T cells. As a result, they are rapidly eliminated by the immune system, forming the basis for clonal deletion [28]. The T-cell immunogenicity of antigens originates from the peptide sequences derived from antigen degradation, known as primary sequences. Through bioinformatics approaches, researchers can now quantitatively analyze the T-cell immunogenicity of antigens [51-54]. Pathogenic microorganisms capable of causing acute infections, such as SARS-CoV-2, possess primary sequences of antigens with highly potent T-cell immunogenicity. This is reflected in our model by larger values of k3 and k10. For such vaccines, the consideration of antigen T-cell immunogenicity is not paramount. However, in the case of chronic infections like HIV, where the T-cell immunogenicity of the antigen is low, it is necessary to moderately enhance the T-cell immunogenicity of the antigen. In immunization, increasing the T-cell immunogenicity of the antigen, represented by the values of k3 and k10, is crucial for boosting the level of neutralizing antibodies. As illustrated in Figure 7a, the IgG antibody level (indicated by the purple dashed line) after secondary immunization with an antigen exhibiting strong T-cell immunogenicity is significantly higher than that achieved with an antigen displaying weak T-cell immunogenicity (indicated by the red dashed line). Figure 7b presents two commonly used methods to enhance the T-cell immunogenicity of antigens, both of which have been extensively applied in practice. The prerequisite for these methods is to not disrupt the antigenic epitopes of the antigen. The first method involves molecular engineering, where antigens are artificially modified without altering their antigenic epitopes. This is achieved by introducing point mutations in internal or non-epitope regions to alter their primary sequences and maximize T-cell immunogenicity. With the advancements in computational protein design technologies, this method has been used to reduce or increase the T-cell immunogenicity of target antigens [55-56]. Another approach is grafting the original antigen onto other proteins to enhance its T-cell immunogenicity. Many vaccines utilize other viral vectors for production, inadvertently leading to the fusion of the target antigen with other protein components. Composite antigens generated through fusion often exhibit stronger T-cell immunogenicity and greatly enhance the induction of neutralizing antibodies. Encouraging results from the clinical trial of the HIV vaccine sv144 demonstrated that increasing the T-cell immunogenicity of antigens has the potential to overcome the challenges of HIV vaccines [57-58]. The RV144 trial, a randomized, double-blind phase 3 efficacy trial, employed a recombinant canarypox vector vaccine, ALVAC-HIV (vCP1521), expressing Env (clade E), group-specific antigen (Gag) (clade B), and protease (Pro) (clade B), along with an alum-adjuvanted AIDSVAX B/E and a bivalent HIV glycoprotein 120 (gp120) subunit vaccine. An underlying risk of this method is that the fusion antigens may introduce additional antigenic epitopes, thereby posing risks of inducing antibody responses against non-target antigenic epitopes.

**Figure 7a:**
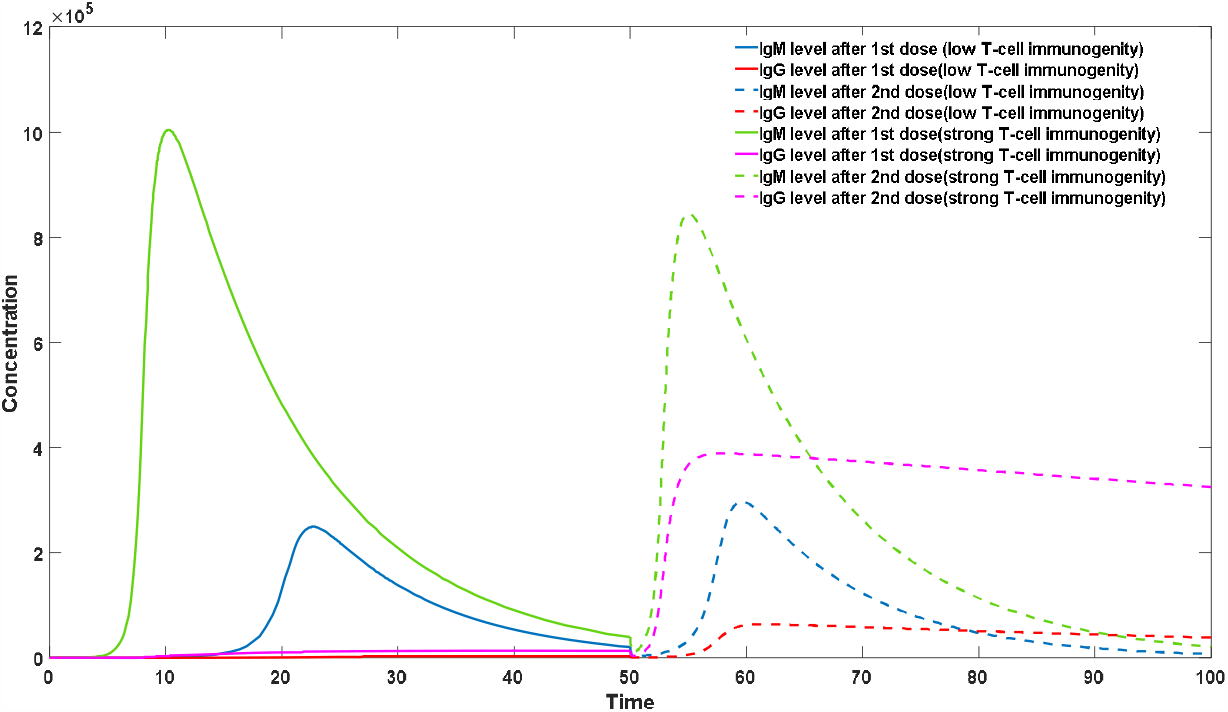
Effect of T-cell immunogenicity on IgG induction. k3 and k10 are assigned to a smaller number (1.5) for the low T-cell immunogenicity group. They are assigned to a larger number (2.5) for its strong T-cell immunogenicity counterpart.

**Figure 7b :**
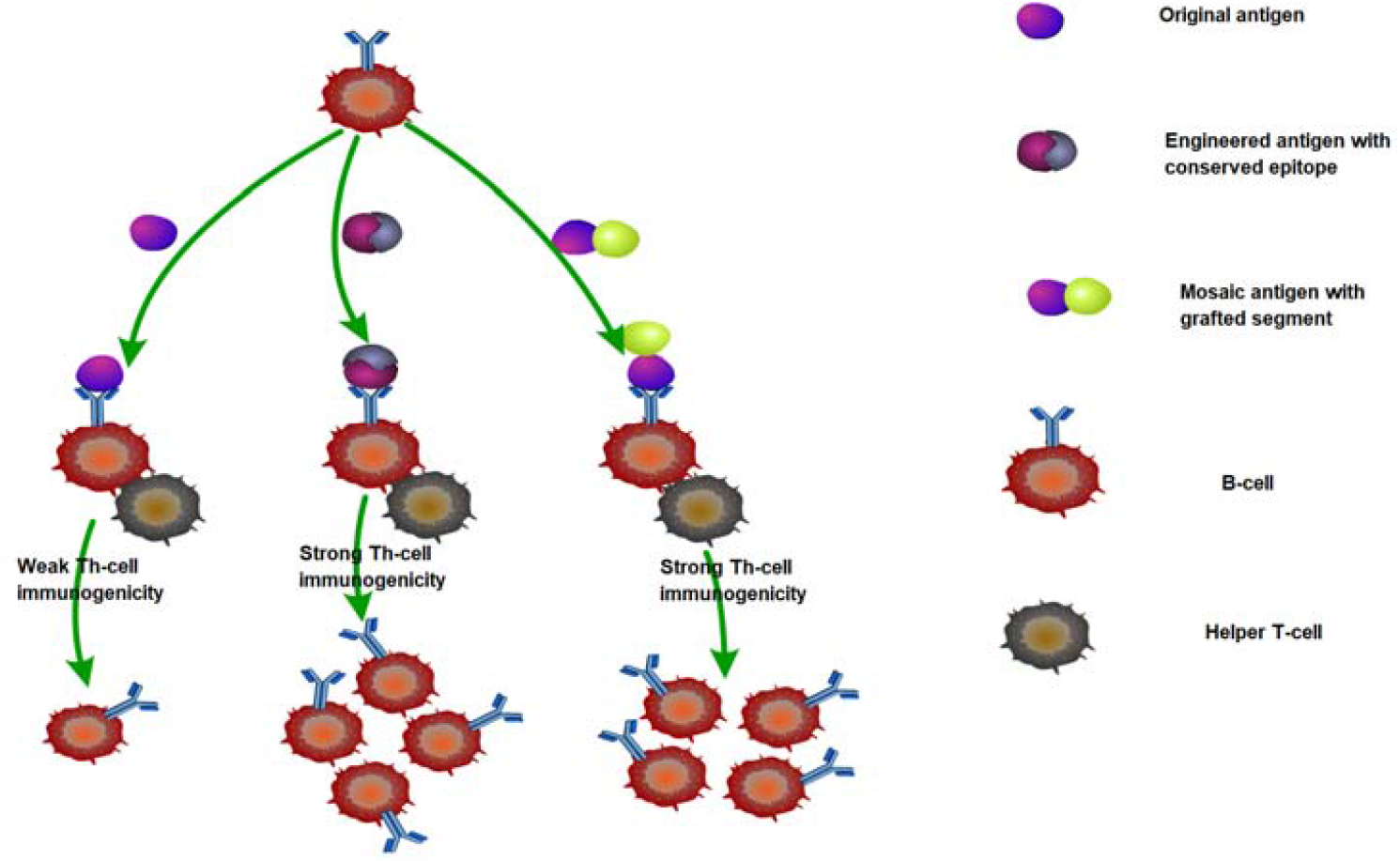
An illustration of approaches in improving Th-cell immonogenity. Two stragties are presented : rational design of antigen with conserved epitope structure and the design of polymer antigen with strong T-cell immonogenity segments.

#### 3.6.2 Directing the Induction of High-affinity Neutralizing Antibodies

Inducing high-affinity antibodies in a targeted manner is a challenging task faced by many vaccine developments. One bottleneck in vaccine development lies in the scarcity of template antibodies in the IgM antibody repertoire that exhibit strong binding to vaccine antigens. This deficiency may stem from the high similarity between the antigenic epitopes of the vaccine antigen and self-antigens, resulting in the loss of highly binding antibodies to the target antigen after clonal deletion. In the case of chronic infections such as HIV, only a small fraction of individuals can generate neutralizing antibodies [59-60]. Directing the induction of neutralizing antibodies can enhance their production levels and likelihood. This directed induction can be achieved through the process depicted in Figure 8a. In 2015, Jardine et al. successfully employed computational protein design techniques to induce mice to produce high concentrations of neutralizing antibodies against the classic HIV antigen, gp120 [61]. The specific approach involved obtaining the crystal structure of the antibody-antigen complex and employing computational protein design methods to introduce point mutations in the antigen’s antibody-binding region, known as the epitope region, to enhance antibody binding affinity. Subsequently, the mutated gp120 was used as the vaccine for the first immunization, followed by a second immunization with the original vaccine. This method yielded superior results compared to traditional two-dose immunization approaches. The simulated process of this method is depicted in Figure 8b. During the simulation, two competitive antibody isotypes were utilized—one with strong binding affinity as a neutralizing antibody and another with weak binding affinity as a non-neutralizing antibody. The initial level of neutralizing antibodies was extremely low at 1e-5, with a high binding capability (k1=1e-5), while the initial level of non-neutralizing antibodies was higher at 1e3 but with lower binding affinity (k1=1e-6). With the employment of the targeted induction approach, the binding affinity between the antigen and antibody was enhanced to 5e-5 due to the altered antigen in the first immunization. However, in the second immunization, when the original antigen was reintroduced, the binding affinity of the target antibodies reverted back to 1e-5. As shown in Figure 8b, the IgG levels induced by the new strategy (represented by the red dashed line) were significantly higher than those induced by the traditional method (represented by the yellow dashed line).

**Figure 8a.**
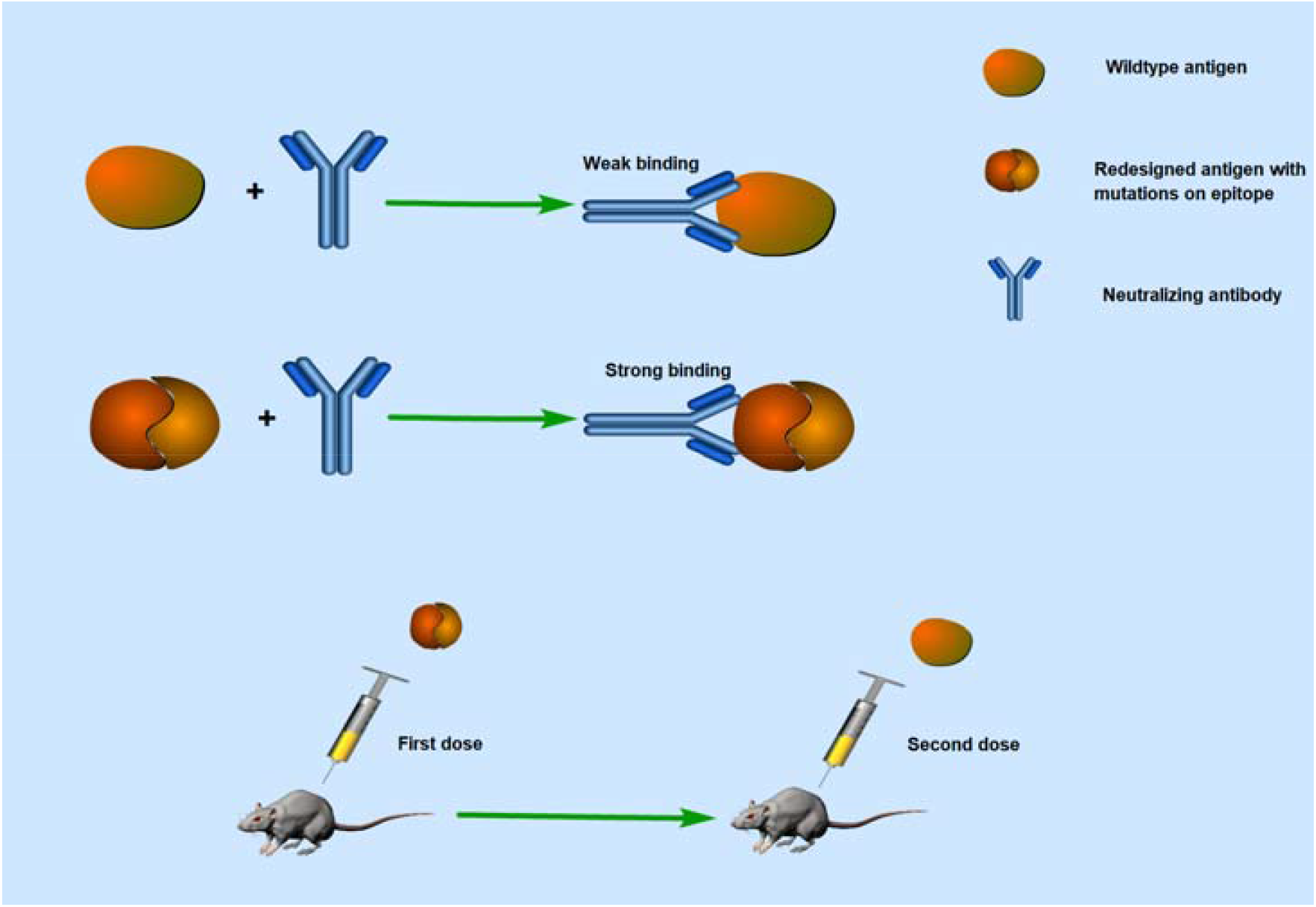
An illustration of induced neutralizing antibody production by mutated antigen. The mutated antigen is used as the vaccine in the first immunization. It was designed to increase the binding affinity with targeted neutralizing antibody.

**Figure 8b.**
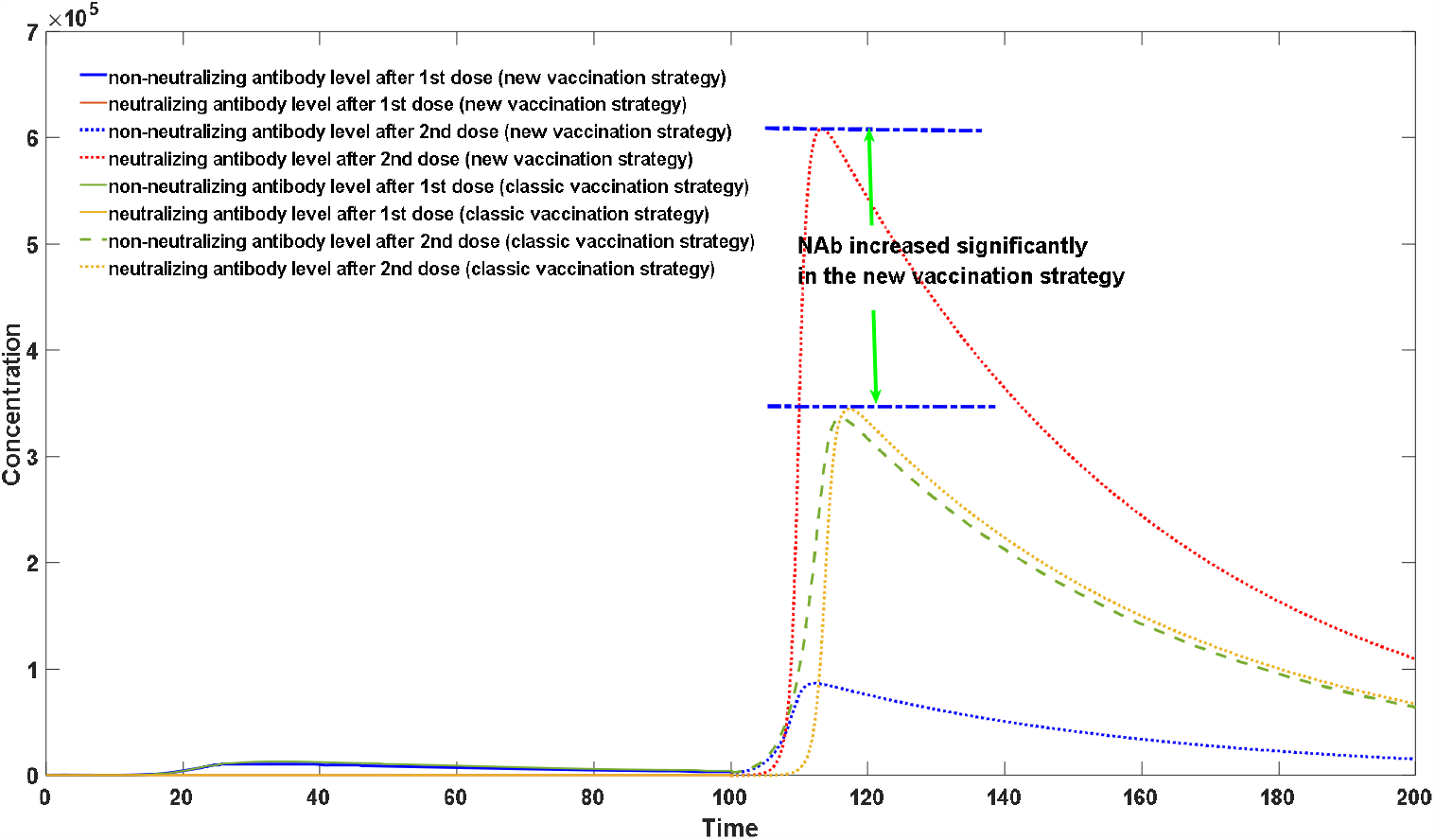
Comparison between induced antibody production and traditional vaccination strategy. First dose is injected at the initial time unit, second dose is injected at 100^th^ time unit. Both injection dosages of antigen substances are 10^6^.

Another more direct approach for inducing the production of neutralizing antibodies is to directly expand the initial levels of antibodies. This method involves the use of gene editing techniques to insert the genes of potent neutralizing antibodies into B cells. Alongside vaccine administration, these genetically edited B cells are injected into the host, resulting in a rapid amplification of neutralizing antibodies [62-64]. This approach is similar to using convalescent blood from recovered individuals to treat certain acute infectious diseases. However, this method may carry significant risks, as these antibodies are not naturally produced by the host and have not undergone the process of clonal selection and deletion, potentially leading to strong immune reactions against host tissues.

#### 3.6.3 Reduce the Decay Rate of IgG

Recurrent viral infections in certain individuals often manifest due to disparate decay rates observed among distinct antibodies, in addition to variations in immunocompetence attributed to factors such as age and overall health. The different decay rate experienced under the complex conditions of self-antigen stimulation contrasts with the conventional direct decay rate of antibodies. Previous investigations have elucidated the pivotal role played by self-antigenic moieties in sustaining IgG levels. The presence of self-antigenic substances precludes a straightforward exponential relationship governing antibody decay, thereby forming the foundation for extended protective periods demonstrated by select antibodies. The critical importance of self-antigens in maintaining antibody titers is exemplified in Figure 9. Case 1 presents the original profile of antibody fluctuations (indicated by the red dashed line). Augmenting the initial concentration of self-antigenic material (Case 2; ranging from 1e5 to 1e6) effectively retards the decline rate of antibodies (depicted by the yellow dashed line). Similarly, amplifying the binding affinity between antibodies and self-antigens (Case 3; increasing from 1e-8 to 5e-8) substantially decelerates antibody decay (shown as the blue dashed line). Notably, while the manipulation of self-antigenic substances remains beyond our purview, we can exert control over antibody attributes. Each antibody variant corresponds to a unique self-antigenic moiety. Certain antibodies exhibit robust self-antigenic moieties, ensuring the maintenance of relatively high concentrations. These antibodies confer prolonged protection against secondary infections, underscoring their significance. Augmenting vaccine-induced protection durations encompasses not only bolstering antibody neutralization capacity but also targeted elicitation of slow-decay-rate antibody responses. It is imperative to recognize that repetitive antigenic exposure solely augments existing antibody levels, without imparting alterations to antibody type and attributes. Consequently, individuals experiencing recurrent infections continue to exhibit antibodies characterized by rapid decay rates, resulting in significantly abbreviated protective cycles relative to their counterparts. This realization underscores that repetitive vaccination does not represent the exclusive optimal strategy.

**Figure 9.**
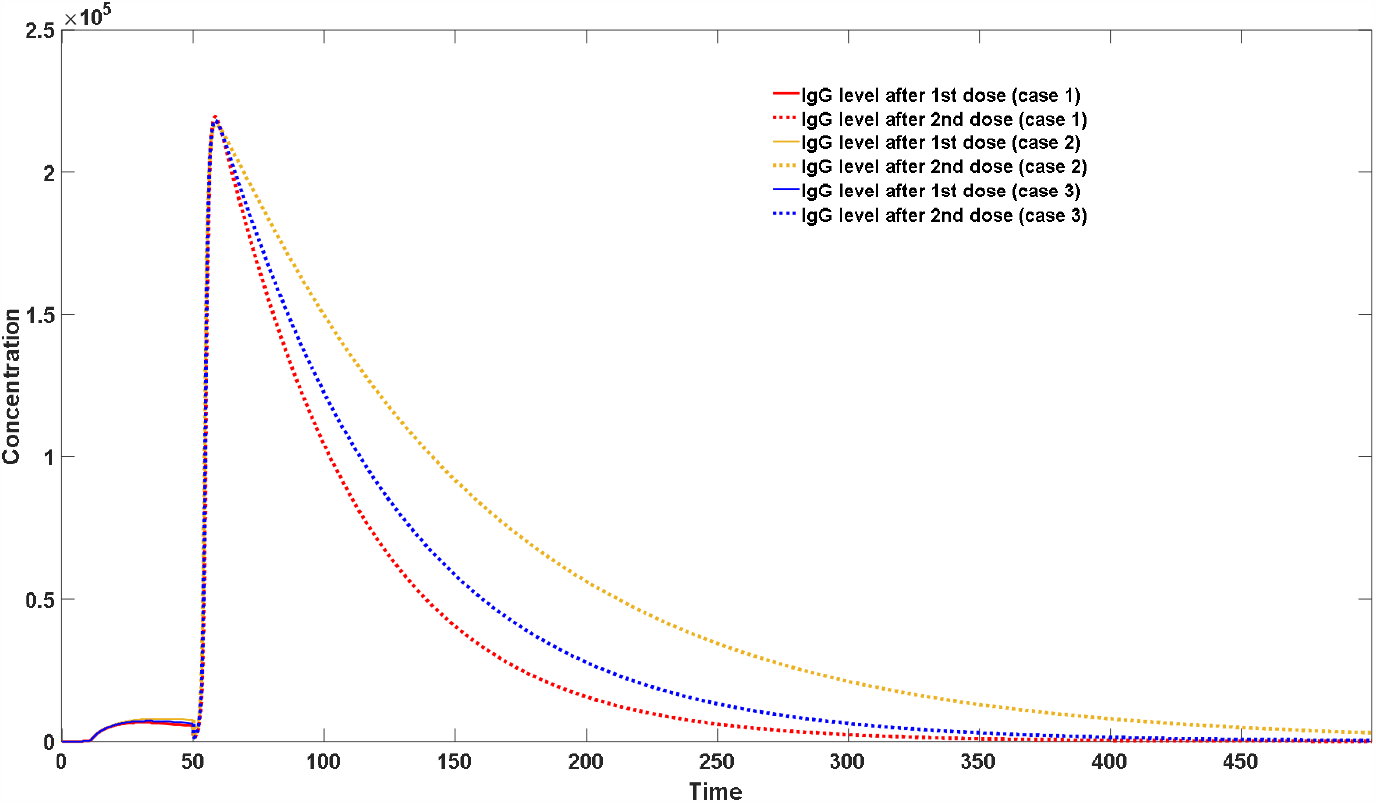
IgG dynamics in different self-antigen scenarios. First dose is injected at the initial time unit, second dose is injected at 50^th^ time unit. Both injection dosages of antigen substances are 10^6^. It can be seen that all IgG would decline after the peak but with different decay speeds.

#### 3.6.4 Reduce the Adverse Effects of Vaccine

All vaccines possess varying degrees of adverse reactions, and we will refrain from delving into the specific adverse effects induced by different vaccines in this context. Instead, our focus lies in introducing the concept that, within our model, adverse reactions can be assessed based on the concentration changes of antigen-antibody complexes. Vaccines function by eliciting the production of IgG, leading to the inevitable generation of complexes with antigenic substances during the antibody synthesis process. While these adverse reactions are unavoidable, the magnitude of adverse effects resulting from vaccines of different types and administration methods at the same antibody induction level exhibits significant variations.

Through systematic comparisons among inactivated vaccines, mRNA vaccines, and attenuated vaccines, we can draw preliminary conclusions. For individuals with normal immune function, achieving equivalent levels of antibody induction, mRNA vaccines and attenuated vaccines yield significantly lower adverse effects compared to inactivated vaccines. In other words, at the same level of adverse reactions, antibodies induced by mRNA vaccines and attenuated vaccines are notably higher than those induced by inactivated vaccines. This observation also elucidates why mRNA vaccines possess superior preventive capabilities against COVID-19 compared to traditional vaccines. Additionally, our model allows for quantitative assessment of such adverse reactions, thereby facilitating better control of vaccine dosages. In the case of mRNA vaccines and inactivated vaccines, adverse reactions demonstrate a significant positive correlation with the administered dosage. Our model provides a theoretical reference for optimal vaccination strategies in future in-silico research.

## 4. Discussion

Studying the interactions between hosts and viruses contributes to understanding the therapeutic mechanisms of infectious diseases and further guides vaccine development. Constructing a rational mathematical model allows for quantitative investigation of the dynamic changes in host-virus interactions, which is advantageous in rational vaccine design. Building upon our previous model of host-virus interactions, we have further developed and refined this model to better simulate the dynamics of antibody changes under different vaccination conditions. Specifically, we have subdivided antibodies into IgM and IgG and incorporated the process of IgM to IgG conversion.

Using this model, we have analyzed the dynamics of antibodies in the body following different vaccine administrations. Specifically, for inactivated vaccines, including protein-based inactivated vaccines produced using exogenous vectors, the IgG levels significantly increase after a secondary dose, thus explaining the importance of sequential vaccination strategies. The initial vaccine dose only activates the production of IgG but does not elevate IgG levels to a higher range. This pattern closely resembles the antibody changes observed during initial infection with blood-borne infectious diseases such as dengue fever virus. Sequential vaccination is also crucial for mRNA vaccines. Compared to traditional inactivated vaccines, mRNA vaccines excel at preserving the original antigenic epitopes. Moreover, due to the gradual release of antigens in mRNA vaccines, they often achieve better antibody enhancement effects while maintaining antigen-antibody complexes at a relatively lower level, resulting in fewer side effects and increased safety. We have further discussed the prospects of attenuated vaccines. We believe that attenuated vaccines hold broader development prospects compared to traditional inactivated vaccines and mRNA vaccines. They do not require specific dosages and can significantly elevate IgG levels without requiring secondary or multiple doses. Other advantages of attenuated vaccines, such as their contagiousness and low side effects, make them a promising choice for future vaccine development.

Based on this model, we have proposed four guiding principles for future vaccine development: increasing the antigen’s T-cell immunogenicity, selectively inducing neutralizing antibodies, selectively inducing antibodies with slow decay rates, and reducing vaccine side effects. These principles are derived from our model-based inferences and have been validated in numerous practical applications.

The development of an HIV vaccine has always been challenging. From a theoretical perspective, we believe there are two core issues. The first issue is the low T-cell immunogenicity of natural antigens, which hinders the effective elevation of antibody levels after vaccine administration. The second issue is the initial low binding activity levels of antibodies in the antibody library, leading to the insufficient increase of neutralizing antibodies after vaccination. To prevent HIV infection, our bodies require antibodies with higher binding affinity to effectively neutralize invading viruses. Once the viruses infiltrate cells, complete clearance becomes difficult, as it is directly related to HIV’s infection of immune cells and the low T-cell immunogenicity. Therefore, to prevent infection, we often need antibodies with stronger binding affinity, known as neutralizing antibodies. Protein engineering can address both of these bottlenecks, especially with the rapid development of computational protein design techniques in recent years. We can enhance the immunogenicity of vaccines by grafting other highly immunogenic proteins and modify the core sequences of antigen proteins without altering the antigenic epitopes, thereby increasing their T-cell immunogenicity. Some groundbreaking work even involves modifying host autologous proteins to stimulate antibody production, which induces immune reactions against self-tissues. This work can also provide valuable insights for future cancer immunotherapy. Through computer-aided protein design, we can design antigens that bind more efficiently to potential neutralizing antibodies, usually achieved through epitope mutations, to selectively enhance their binding affinity. Of course, this technique may involve experimental approaches such as protein-directed evolution. Using engineered antigens as the initial immunogenic substances for primary vaccination, followed by secondary vaccination using the original antigens, can induce higher levels of neutralizing antibodies. By efficiently combining these two methods, we may achieve greater breakthroughs in HIV vaccine research.

Our model indicates that the decay effects of antibodies may vary significantly among individuals due to their inherent properties. Some antibodies exhibit good self-immunogenicity and can sustain at a high level, leading to long-lasting protection. Conversely, some antibodies demonstrate lower self-immunogenicity and exhibit faster decay rates without stimulation from self-antigen substances, resulting in a higher risk of recurrent infections. In particular those might due to the composition of the individual innate repertoire. This original antibody composition comes partly from the scarcity of IgMs 2 the innate repertoire mentioned in Subsection 3.6., which comes from the limited size of the V(D)J DNA sequences which are their source in chromosome 14 [65]. Indeed, the genes coding for IgMs are located on chromosome 14 for the chain heavy and on chromosomes 2 and 22 for the alternative light chain loci, (kappa and lambda, respectively). The process of DNA recombination in bone marrow B lymphocytes generates a diverse range of immunoglobulins through the random recombination of different DNA segments of the V(D)J complex [66-68], whose composition is hereditary and whose heterogeneity is explained by the selection to which it has been subjected by contact with lethal infectious agents during human evolution. This heterogeneity is not taken into account in this model, but could be in a later version incorporating the genetic regulation of the immune system [69]. Besides individual differences, age and health factors also influence the ability to stimulate antibody regeneration by T-cells. Healthy individuals tend to generate more antibodies under the same conditions of self-antigen substance, leading to a decrease in the antibody decay rate. As a result, individuals with compromised immune systems are more prone to recurrent infections, such as COVID-19. Our model also suggests that repeated administration of the same vaccine is not conducive to altering the original antibody composition. Therefore, we do not consider repetitive vaccination as a long-term solution for preventing infections. Future vaccine development, such as for COVID-19, should focus on selectively inducing high-binding and slow-decay antibodies. Furthermore, our model quantitatively reflects the concentration of antigen-antibody complexes, which can be used as an indicator of vaccine side effects, including the severity of symptoms after natural infection. Our research demonstrates that different vaccines may exhibit significant differences in terms of inducing the same level of IgG. Thus, selecting vaccines with minimal side effects is an important consideration for future vaccine development.

Finally, our theoretical research also lays a foundation for future endeavors pertaining to the development of vaccines based on mathematical modeling. Nevertheless, it is imperative to recognize a fundamental premise: that all models, by their very nature, are fallible, albeit they possess varying degrees of utility. It is evident that the immune system possesses a remarkably intricate and heterogeneous nature. Regrettably, we have allocated limited attention to the facets of innate immunity such as the production of interferon regulated by a complex genetic network [69], primarily directing our scientific inquiries towards humoral immunity. We also ignored the important roles of somatic hypermutation on the generation of neutralizing antibody and the gradual loss of antibody binding capacity after infection [28]. It is important to acknowledge the inherent uncertainties associated with our model, and its refinement will necessitate ongoing validation through additional experimental investigations and clinical data.

## Data Availability

All supplementary materials and Matlab codes can be accessed at: https://github.com/zhaobinxu23/vaccine_modeling

## Author Contributions

Conceptualization, Z.X. and J.D.; methodology, Z.X.; writing—original draft preparation, Z.X. and S.J.; writing—review and editing, D.W., H.Z. and J.D.; funding acquisition, Z.X. All authors have read and agreed to the published version of the manuscript.

## Funding

This research was funded by DeZhou University, grant number 30101418.

## Acknowledgments

We thank Dr.Zuyi Huang from Villanova University. Dr.Yushan Zhu from Tsinghua University, Peiyan Guan, XiangYong Li, and Tianxiang Chen from Dezhou University for helpful conversations, comments, and clarifications.

## Conflicts of Interest

The authors declare no conflict of interest.

## Notes

### Competing Interest Statement

The authors have declared no competing interest.

